# Clinical characteristics of 101 COVID-19 nonsurvivors in Wuhan, China: a retrospective study

**DOI:** 10.1101/2020.03.04.20031039

**Authors:** Qiao Shi, Kailiang Zhao, Jia Yu, Fang Jiang, Jiarui Feng, Kaiping Zhao, Xiaoyi Zhang, Xiaoyan Chen, Peng Hu, Yupu Hong, Man Li, Fang Liu, Chen Chen, Weixing Wang

## Abstract

**Background:** The outbreak of COVID-19 has aroused global concerns. We aimed to describe the clinical characteristics of COVID-19 nonsurvivors and analyze possible causes for the rapid disease progress to death.

**Methods:** Patients with confirmed COVID-19 died in Renmin Hospital of Wuhan University before February 15, 2020, were identified. We obtained epidemiological, demographic, and clinical data from electronic medical records.

**Results:** Among 101 nonsurvivors, the median age was 71.0 years (IQR, 59.0-80.0), 59.4% were male, 79.2% had one or more comorbidities including hypertension (58.4%), cardiovascular disease (22.8%), diabetes (20.8%) etc. The most common symptoms were fever (76.2%), cough (58.4%) and dyspnea (54.5%). Respiratory failure (99.0%), acute cardiac injury (52.5%), sepsis (40.6%) and acute kidney injury (23.8%) were most common complications. Compared with patients died after 3 days of admission, patients died within 3 days of admission had significantly higher white blood cell count (10.8 vs 6.7×10^9^/L, *P*=0.001) and neutrophil count (8.9 vs 5.5×10^9^/L, *P*=0.001), longer prothrombin time (13.2 vs 12.5 s, *P*=0.040), higher D-dimer concentration (7.64 vs 2.82, *P*=0.040), higher lactate level (2.9 vs 2.2 mmol/L, *P*=0.042), lower oxygen saturation (85.0% vs 91.0%, *P*=0.008), and were more likely to suffer sepsis (52.1% vs 30.2%, *P*=0.025).

**Conclusions:** Older patients with underlying comorbidities suffering COVID-19 were at high risk of death. Respiratory failure, acute cardiac injury and acute kidney injury played crucial roles in the death of COVID-19 patients. Early development of sepsis was associated with the rapid disease progress to death.

## INTRODUCTION

Since December 2019, the outbreak of coronavirus disease 2019 (COVID-19) caused by severe acute respiratory syndrome coronavirus 2 (SARS-CoV-2) swept across China and many other countries have caused global concerns (1). As May 4, 2020, a total of 3, 517, 345 COVID-19 cases around the world have been confirmed and 243, 401 lost their lives (2). In the early outbreak of COVID-19, respiratory infection symptoms occurred, with some patients rapidly developed acute respiratory distress syndrome (ARDS), acute respiratory failure and even death. Compared with severe acute respiratory syndrome (SARS) and Middle East respiratory syndrome (MERS), the concealed onset, stronger contagion and relatively lower mortality could be defined as the clinical features of the disease (3-5). However, many patients suffered rapid disease progress to death after admission in the early breakout in Wuhan, China.

Previous studies focused on clinical characteristics and outcomes of COVID-19 patients varied from mild, severe, even deceased cases (6,7). But the possible causes for the rapid disease progress to death need to be addressed further. In this study, we aimed to describe the clinical characteristics and outcomes of nonsurvivors confirmed with SARS-CoV-2 infection in the early outbreak, and to analyze the possible causes for the rapid disease progress to death after admission in patients with COVID-19.

## METHODS

### Study Design and Participants

All the patients with confirmed COVID-19 hospitalized in Renmin Hospital of Wuhan University (including Shouyi campus and Eastern campus) died before February 15, 2020 in Wuhan, China, were enrolled. Renmin Hospital of Wuhan University is one of the major tertiary hospitals in Wuhan, China and is a government designated hospital for COVID-19 treatment. All patients with COVID-19 identified in this study were diagnosed according to World Health Organization interim guidance (8). Large proportion of patients died within 3 days of admission in the early outbreak of COVID-19, we also compared demographic and clinical characteristics of patients died within 3 days with those survived 3 days of admission.

### Procedures

We obtained epidemiological, demographic, laboratory, and outcome data from electronic medical records. Missing data were collected through direct communication with involved health-care providers or family member of patients. All data were checked by two sophisticated physicians.

The date of disease onset was defined as the day when the onset of symptoms were noticed. Acute respiratory distress syndrome (ARDS) and acute respiratory failure were defined according to the Berlin definition and previously reported (9,10). Acute kidney injury (AKI) was identified based on serum creatinine (11). Acute liver injury was identified according to previous report (12). Acute cardiac injury was defined if the serum ultra-sensitive troponin I was above the 99th percentile upper reference limit (6). Sepsis was defined according to the Third International Consensus Definitions for Sepsis and Septic Shock (13). Characteristic chest computer tomography (CT) imaging of these nonsurvivors were evaluated by previously described method (14).

The primary outcome was the possible causes for the rapid disease progress to death after admission. Secondary outcomes were to present epidemiological and demographic data, comorbidities, signs and symptoms on admission, laboratory indicators, CT findings, treatments, organ injury or failure (including lung, heart, kidney and liver etc.), and to compare the differences of patients died within 3 days with those survived 3 days of admission.

### Statistical analysis

Continuous variables were described as the medians and interquartile ranges (IQRs). Categorical variables were described as frequencies and percentages based on the available data. We used Pearson’s chi-square test, the Mann-Whitney test and Fisher’s exact test for comparisons between patients died within 3 days and those died after 3 days of admission, as appropriate. Survival curve was developed using the Kaplan-Meier method with log-rank test. All statistical analyses were performed using SPSS (version 22.0) and GraphPad Prism (version 5.0) software. *P*<0.05 was considered statistically significant.

## RESULTS

By February 15, 2020, 101 hospitalised patients with confirmed COVID-19 died in Renmin Hospital of Wuhan University (including Shouyi campus and Eastern campus) were included in this study. The median age of nonsurvivors was 71.0 (IQR, 59.0-80.0) years, 59.4% were male. There were 57 (56.4%) cases whose age over 70 years (Table 1). Among all the nonsurvivors, 81 (79.2%) patients had one or more underling medical conditions. The most common comorbidities included hypertension (58.4%), cardiovascular disease (22.8%), diabetes (20.8%), chronic pulmonary disease (14.9%), and cerebrovascular disease (12.9%). The most common symptoms were fever (76.2%), cough (58.4%), dyspnea (54.5%), and fatigue (37.6%). Other less common symptoms were diarrhea (9.9%), anorexia (5.9%), nausea and vomiting (2.0%) and abdominal pain (1.0%) (Table 1). The median duration from the onset of symptoms to admission and from admission to death were 9.0 (IQR, 5.0-13.0) days and 4.0 days (IQR, 2.0-7.0), respectively. The survival curve of COVID-19 patients were shown in Figure 1.

**Table 1.** Characteristics of COVID-19 nonsurvivors.

| **Variables** | **Total (N=101)** | **Patients died within 3 days (N=48)** | **Patients survived 3 days (N=53)** | **P Value\*** |
| --- | --- | --- | --- | --- |
| **Age, years** | 71.0 (59.0-80.0) | 72.0 (59.0-78.0) | 71.0 (59.0-81.0) | 0.838 |
| < 70 years | 44 (43.6) | 14 (39.6) | 17 (47.2) | 0.443 |
| ≥ 70 years | 57 (56.4) | 34 (60.4) | 36 (52.8) |  |
| Sex, male | 60 (59.4) | 28 (58.3) | 32 (60.4) | 0.835 |
| Smoking | 5 (5.0) | 2 (4.2) | 3 (5.7) | 1.000 |
| Drinking | 5 (5.0) | 1 (2.1) | 4 (7.5) | 0.365 |
| Exposure history | 4 (4.0) | 3 (6.3) | 1 (1.9) | 0.344 |
| **Comorbidities** |  |  |  |  |
| Hypertension | 59 (58.4) | 27 (56.3) | 32 (60.4) | 0.674 |
| Cardiovascular disease | 23 (22.8) | 9 (18.8) | 14 (26.4) | 0.359 |
| Diabetes | 21 (20.8) | 9 (18.8) | 12 (22.6) | 0.630 |
| Chronic pulmonary disease | 15 (14.9) | 6 (12.5) | 9 (17.0) | 0.527 |
| Cerebrovascular disease | 13 (12.9) | 4 (8.3) | 9 (17.0) | 0.195 |
| Chronic kidney disease | 10 (9.9) | 2 (4.2) | 8 (15.1) | 0.096 |
| Chronic liver disease | 5 (5.0) | 1 (2.1) | 4 (7.5) | 0.365 |
| Malignancy | 7 (6.9) | 1 (2.1) | 6 (11.3) | 0.115 |
| **Number of comorbidities** |  |  |  | 0.132 |
| 0 | 2 (21.8) | 12 (25.0) | 10 (18.9) | .. |
| 1 | 37 (36.6) | 21 (43.8) | 16 (30.2) | .. |
| ≥2 | 42 (41.6) | 15 (31.3) | 27 (50.9) | .. |
| **Signs and Symptoms** |  |  |  |  |
| Fever | 77 (76.2) | 36 (75.0) | 41 (77.4) | 0.781 |
| Dyspnea | 55 (54.5) | 27 (56.3) | 28 (52.8) | 0.730 |
| Dry cough | 59 (58.4) | 26 (54.2) | 33 (62.3) | 0.410 |
| Fatigue | 38 (37.6) | 20 (41.7) | 18 (34.0) | 0.425 |
| Diarrhea | 10 (9.9) | 4 (8.3) | 6 (11.3) | 0.744 |
| Anorexia | 6 (5.9) | 3 (6.3) | 3 (5.7) | 1.000 |
| Abdominal pain | 1 (1.0) | 0 | 1 (1.9) | 1.000 |
| Nausea or vomiting | 2 (2.0) | 1 (2.1) | 1 (1.9) | 1.000 |
| **Vital signs on admission, Median (IQR)** |  |  |  |  |
| HR, bpm | 88 (78-105) | 97 (80-112) | 84 (74-95) | **0.006** |
| RR, rpm | 21 (19-30) | 25 (20-36) | 20 (19-24) | **0.005** |
| MAP, mmHg | 93 (83-105) | 93 (81-107) | 93 (85-105) | 0.859 |
| Lung severity score | 14 (6-18) | 13 (6-18) | 14 (5-19) | 0.899 |
| From symptoms to admission, days | 9.0 (5.0-13.0) | 10.0 (6.0-13.0) | 8.0 (4.0-14.0) | 0.472 |
| From admission to death, days | 4.0 (2.0-7.0) | 2.0 (1.0-3.0) | 7.0 (5.0-8.0) | **<0.001** |

Data are expressed as median (IQR) or n (%). \*P values indicate differences between patients died within 3 days of admission and those survived 3 days of admission. $P < 0.05$ was considered statistically significant. Abbreviations: HR= heart rate; IQR= interquartile range; MAP= mean arterial pressure; RR= respiratory rate.

**Figure 1.**
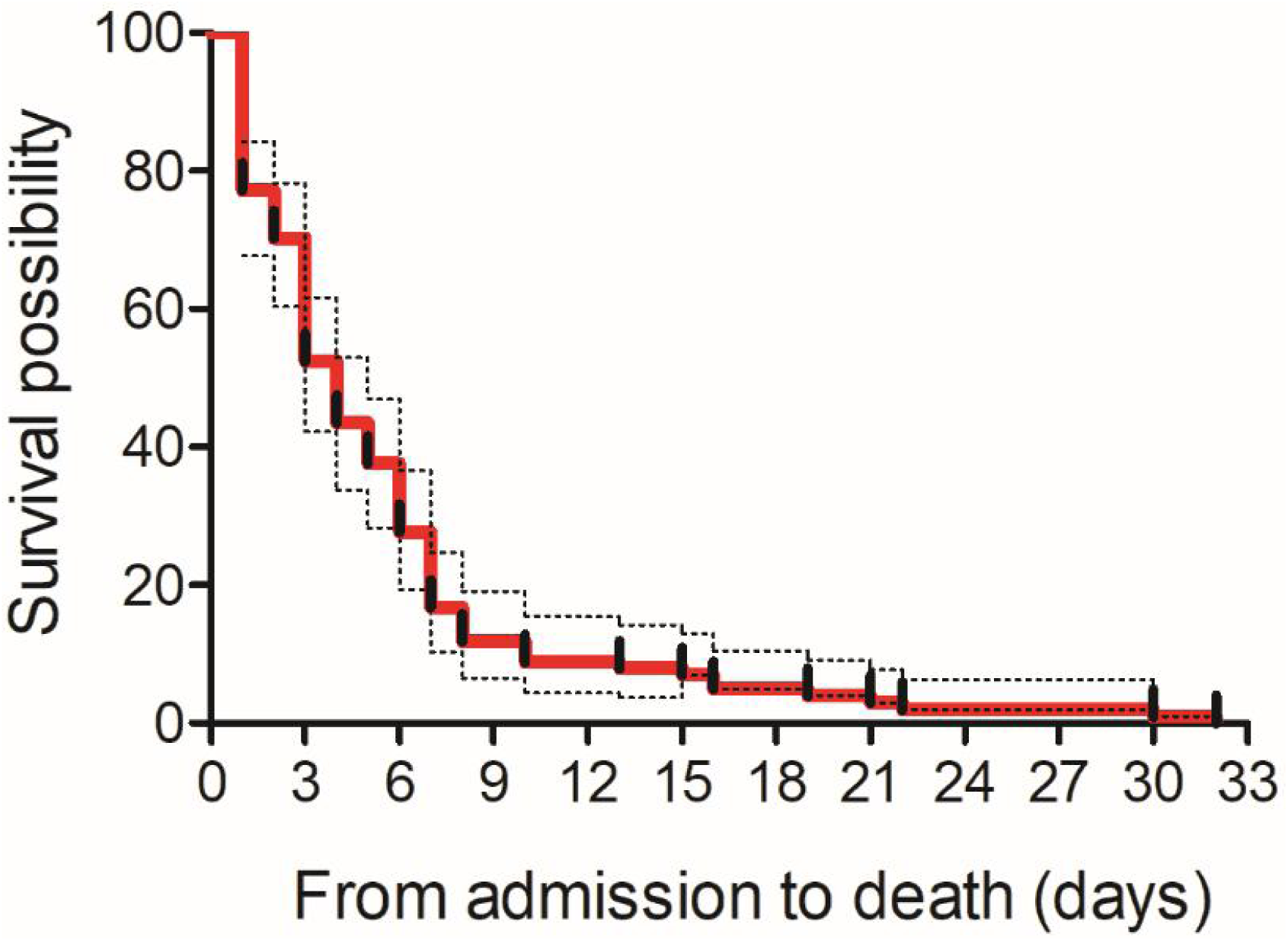
Survival curve of COVID-19 nonsurvivors.

Forty-eight cases (47.52%) died within 3 days of admission, the median age was 72.0 (IQR, 59.0-78.0) years, and 58.3% were male. Fifty-three cases (52.5%) died after 3 days of admission, the median age was 71.0 (IQR, 59.0-81.0) years, and 60.4% were male. Compared with patients who survived 3 days, those died within 3 days of admission exhibited significantly higher respiration rate (25 *vs* 20 rpm, *P*=0.005) and heart rate (97 *vs* 84 bpm, *P*=0.006) on admission. There were no significant differences of self-report symptoms and underlying comorbidities between patients died within 3 days of admission and those survived 3 days of admission (Table 1).

We observed substantial abnormalities (below or over normal range) in laboratory findings of these nonsurvivors on admission (Table 2). Elevated neutrophil count, increased levels of C-reactive protein, procalcitonin, NT-proBNP, ultra-sensitivity troponin I, myoglobin, blood glucose, D-dimer and lactate were noticed in the nonsurvivors. Besides, significantly decreased lymphocyte count, PaO_2_ and SpO_2_ below normal range were also obvious in these nonsurvivors. Patients died within 3 days of admission had significantly higher white blood cell count (10.8 *vs* 6.7×10^9^/L, *P*= 0.001), neutrophil count (8.9 *vs* 5.5×10^9^/L, *P*=0.001). For other biochemical indexes, creatine kinase-MB (3.3 *vs* 2.0 ng/ml, *P*=0.036), myoglobin (186.6 *vs* 118.7 ug/L, *P*=0.034) were significantly higher in patients died within 3 days of admission. Compared with patients survived 3 days of admission, those died within 3 days had longer prothrombin time (13.2 *vs* 12.5 s, *P*=0.040) and higher D-dimer concentration (7.64 *vs* 2.82, *P*=0.040). Blood gas analyses showed patients died within 3 days presented with significantly lower oxygen saturation (85.0% *vs* 91.0%, *P*=0.008) and higher lactate level (2.9 *vs* 2.2 mmol/L, *P*=0.042) which reflected poor oxygenation in these patients. Patients suffered severe lung damage and their lung CT features presented vast or merged glass shadow, but quantification of lung injury revealed no significant difference between patients died within 3 days and those died after 3 days of admission (Table 1 and Figure 2).

**Table 2.** Laboratory findings of COVID-19 nonsurvivors on admission.

| Variables | Normal Range | Total (N=101) | Patients died within 3 days (N=48) | Patients survived 3 days (N=53) | P Value* |
| --- | --- | --- | --- | --- | --- |
| White blood cell count, $\times 10^9/L$ | 3.5-9.5 | 8.2 (5.6-12.9) | 10.8 (8.0-14.0) | 6.7 (4.8-10.9) | **0.001** |
| Neutrophil count, $\times 10^9/L$ | 1.8-6.3 | 7.1 (4.4-11.5) | 8.9 (6.6-12.8) | 5.5 (3.8-8.9) | **0.001** |
| Lymphocyte count, $\times 10^9/L$ | 1.1-3.2 | 0.6 (0.4-0.9) | 0.6 (0.4-0.9) | 0.6 (0.4-0.8) | 0.851 |
| Platelet count, $\times 10^9/L$ | 125-350 | 157 (118-212) | 164 (105-239) | 157 (122-203) | 0.983 |
| C-reactive protein, mg/L | 0.068-8.2 | 105.9 (58.4-173.4) | 119.8 (65.5-186.6) | 86.7 (55.2-141.1) | 0.094 |
| Procalcitonin, ng/ml | <0.1 | 0.3 (0.1-1.4) | 0.5 (0.1-2.4) | 0.2 (0.1-0.5) | 0.060 |
| NT-proBNP, pg/mL | 0-400 | 838 (351-2029) | 889 (330-2728) | 817 (358-1701) | 0.608 |
| Ultra-sensitivity troponin I, ng/ml | 0-0.04 | 0.05 (0.02-0.17) | 0.06 (0.02-1.33) | 0.05 (0.02-0.11) | 0.139 |
| Creatine kinase-MB, ng/ml | 0-5 | 2.6 (1.2-4.5) | 3.3 (2.1-7.7) | 2.0 (1.0-4.1) | **0.036** |
| Myoglobin, ug/L | 0-110 | 125.0 (73.9-473.8) | 186.6 (84.6-1000.0) | 118.7 (64.1-344.7) | **0.034** |
| Alanine aminotransferase, U/L | 7-40 | 25.0 (18.0-48.3) | 27.5 (18.0-54.8) | 23.0 (16.3-41.5) | 0.111 |
| Aspartate aminotransferase, U/L | 13-35 | 41.5 (29.0-64.5) | 50.5 (29.3-73.8) | 39.0 (28.0-52.0) | 0.096 |
| Albumin, g/L | 40-55 | 33.0 (30.3-36.4) | 32.5 (29.1-35.1) | 33.1 (30.9-37.0) | 0.225 |
| Total bilirubin, mmol/L | 0-23 | 12.0 (8.4-19.1) | 12.2 (8.9-21.8) | 12.0 (8.1-17.5) | 0.611 |
| Blood urea nitrogen, mmol/L | 3.1-8.8 | 8.5 (5.5-14.4) | 9.3 (6.8-19.9) | 7.3 (5.4-13.5) | 0.066 |
| Creatinine, $\mu\text{mol/L}$ | 41-81 | 79.0 (53.5-128.0) | 84.0 (55.8-154.0) | 73.0 (53.0-116.8) | 0.473 |
| Blood glucose, mmol/L | 3.9-6.1 | 7.3 (5.9-9.4) | 7.3 (6.2-9.5) | 7.3 (5.6-9.4) | 0.484 |
| Prothrombin time, s | 9-13 | 12.8 (12.0-14.0) | 13.2 (12.5-14.4) | 12.5 (11.7-13.8) | **0.040** |
| Activated partial thromboplastin time, s | 25.1-36.5 | 29.6 (27.9-33.3) | 29.8 (28.6-33.7) | 29.6 (27.7-33.2) | 0.638 |
| D-dimer, mg/L | 0-0.55 | 3.95 (0.98-14.82) | 7.64 (2.31-17.99) | 2.82 (0.76-13.12) | **0.040** |
| CD3<sup>+</sup> cell count, /ul | 723-2737 | 306.0 (184.8-479.8) | 252.5 (180.5-403.0) | 326.5 (230.0-487.0) | 0.356 |
| CD4<sup>+</sup> cell count, /ul | 404-1612 | 185.0 (120.3-281.5) | 180.0 (126.0-298.0) | 188.5 (106.8-278.5) | 0.744 |
| CD8<sup>+</sup> cell count, /ul | 220-1129 | 111.0 (52.0-158.3) | 70.0 (47.3-150.0) | 132.0 (61.8-168.0) | 0.133 |
| **Blood gas analysis** |  |  |  |  |  |
| pH | 7.35-7.45 | 7.42 (7.35-7.46) | 7.40 (7.29-7.45) | 7.43 (7.39-7.47) | 0.057 |
| PaO<sub>2</sub>, mmHg | 80-100 | 54.5 (41.0-69.8) | 49.5 (37.3-65.0) | 57.0 (45.8-80.5) | 0.063 |
| PaCO<sub>2</sub>, mmHg | 35-45 | 36.5 (30.8-41.0) | 38.0 (29.3-42.0) | 34.5 (31.0-40.3) | 0.364 |
| SpO<sub>2</sub>, % | 95-98 | 89.0 (77.8-94.0) | 85.0 (71.5-92.5) | 91.0 (81.5-96.0) | **0.008** |
| Lactate, mmol/L | 0.5-1.5 | 2.6 (1.9-3.6) | 2.9 (2.2-4.2) | 2.2 (1.4-3.6) | **0.042** |

Data are expressed as median (IQR). \*P values indicate differences between patients died within 3 days of admission and those survived 3 days of admission. $P < 0.05$ was considered statistically significant. Abbreviations: PaCO<sub>2</sub>=partial pressure of carbon dioxide; PaO<sub>2</sub>= partial pressure of oxygen; SpO<sub>2</sub>= oxygen saturation.

**Figure 2.**
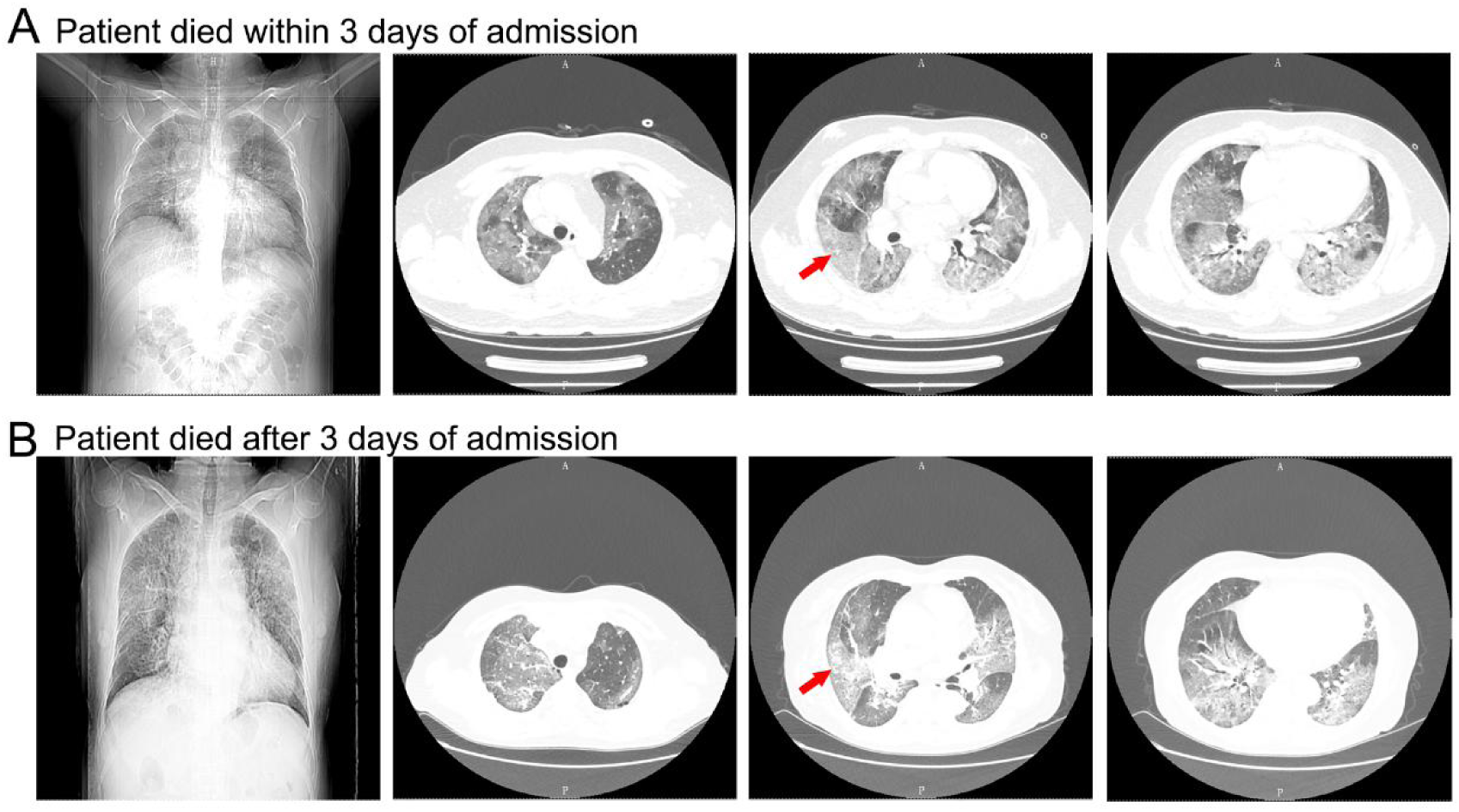
Chest CT manifestations of COVID-19 nonsurvivors. A: Chest computed tomographic images obtained from a 47-year-old male patient on day 6 after onset of symptoms showed extensive and fused ground glass opacity (red arrow) in both lungs. B: Chest computed tomographic images obtained from a 59-year-old female on day 7 after onset of symptoms showed extensive and fused ground glass opacity (red arrow) in both lungs.

Among treatment strategies, most patients received antiviral therapy (96.0%) and antibacterial therapy (98.0%). Many received glucocorticoid therapy (43.6%) and immunoglobulin therapy (35.6%). Seventy-eight (77.2%) patients received high flow oxygen inhalation, 40.6% received noninvasive mechanical ventilation while 13.9% received invasive mechanical ventilation and 5.0% received continuous renal replacement therapy (Table 3).

**Table 3.** Complications and treatments of COVID-19 nonsurvivors.

| **Variables** | **Total (N=101)** | **Patients died within**<br>**3 days (N=48)** | **Patients survived**<br>**3 days (N=53)** | **P Value\*** |
| --- | --- | --- | --- | --- |
| **Treatments** |  |  |  |  |
| Antiviral therapy | 97 (96.0) | 47 (97.9) | 50 (94.3) | 0.619 |
| Antibiotic therapy | 99 (98.0) | 47 (97.9) | 52 (98.1) | 1.000 |
| Glucocorticoid therapy | 44 (43.6) | 17 (35.4) | 27 (50.9) | 0.116 |
| Immunoglobulin therapy | 36 (35.6) | 13 (27.1) | 23 (43.4) | 0.087 |
| High flow oxygen inhalation | 78 (77.2) | 35 (72.9) | 43 (81.1) | 0.326 |
| NIMV | 41 (40.6) | 11 (22.9) | 30 (56.6) | **0.001** |
| IMV | 14 (13.9) | 8 (16.7) | 6 (11.3) | 0.437 |
| CRRT | 5 (5.0) | 2 (4.2) | 3 (5.7) | 1.000 |
| **Complications** |  |  |  |  |
| Respiratory failure | 100 (99.0) | 47 (97.9) | 53 (100.0) | 0.475 |
| Acute cardiac injury | 53 (52.5) | 28 (58.3) | 25 (47.2) | 0.262 |
| Acute kidney injury | 24 (23.8) | 12 (25.0) | 12 (22.6) | 0.781 |
| Acute liver injury | 18 (17.8) | 9 (18.8) | 9 (17.0) | 0.817 |
| Coagulation disorders | 15 (14.9) | 4 (8.3) | 11 (20.8) | 0.080 |
| Sepsis | 41 (40.6) | 25 (52.1) | 16 (30.2) | **0.025** |

Data are expressed as n (%). \**P* values indicate differences between patients died within 3 days of admission and those survived 3 days of admission. *P*<0.05 was considered statistically significant. Abbreviations: CRRT=continuous renal replacement therapy; IMV= invasive mechanical ventilation; NIMV= noninvasive mechanical ventilation.

Regarding to organ functions, 100 (99.0%) patients developed respiratory failure, 52.5% had acute cardiac injury, 40.6% developed sepsis, acute kidney injury were developed in 24 cases (23.8%), acute liver dysfunction were found in 18 cases (17.8%) and 15 (14.9%) suffered coagulation disorders among which one patient died of massive upper gastrointestinal bleeding. Compared with patients died after 3 days of admission, patients died within 3 days of admission were more likely to develop sepsis. (52.1% *vs* 30.2%, *P*=0.025) (Table 3).

## DISUSSION

As of February 15, 2020, 101 patients died because of COVID-19 were included in this study. In the early outbreak of COVID-19, major gaps in our knowledge of the epidemiology, clinical features and treatment measures of COVID-19 posed huge challenge for our clinical practice. Although previous study reported the mortality of patients with COVID-19 was 1.4% (15), some special population including the aged and individuals with underlying comorbidities may be vulnerable to SARS-CoV-2 infection. Recent publication has demonstrated old age as an independent predictor of mortality in COVID-19 patients (16). In our study, the median age of nonsurvivors was 71.0 years, 79.2% of them combined with one or more underlying medical conditions including hypertension, cardiovascular disease, diabetes and chronic pulmonary disease etc. This suggested that aged individuals with chronic medical conditions were more likely to suffer a severe attack and death.

Common symptoms at onset of COVID-19 were fever, dry cough, myalgia, fatigue etc. (6,17). In our study, we observed that besides fever and cough, dyspnea presented early in most nonsurvivors with confirmed COVID-19, similar finding has also been observed in critically ill patients reported by Yang etc. (18). Recent publication suggested the median time from onset of symptoms to hospital admission of deceased patients was 10.0 (IQR, 7.0-13.0) days, which tended to be longer than that of recovered patients (9.0 [IQR, 6.0-12.0] days) (19). In our study, the median time from symptom onset to admission of patients died within 3 days and after 3 days of admission was 10.0 (IQR, 6.0-13.0) days and 8.0 (IQR, 4.0-14.0) days, respectively, but with no significantly difference between two groups. This indicated that although longer duration before admission might be associated with poor prognosis of COVID-19 patients, it was not contributed to the rapid disease progress to death within 3 days of admission.

Abnormal laboratory findings on admission suggested impaired pulmonary, cardiac, liver and coagulation functions as well as disturbed immune function of these nonsurvivors even before hospitalisation. ARDS was the most common complication in nonsurviving patients with COVID-19 and oxygen saturation was an important manifestation of ARDS. In our study, SpO_2_ of patients died within 3 days of admission was 85.0% on admission, which was significantly lower compared with those died after 3 days of admission. These suggested that SpO_2_ on admission might serve as early warning signals of poor prognosis and should attract more attentions. In terms of SARS, severe respiratory failure was the leading cause of mortality, with little other organ failure (20). In our study, it was noticed that 52.5% and 23.8% nonsurvivors suffered acute cardiac injury and AKI, respectively. Angiotensin-converting enzyme 2 (ACE2), a crucial SARS-CoV-2 receptor, distributed widely in heart, kidney, and intestine etc. (21-23). Whether SARS-CoV-2 attack heart and kidney or other vital organs directly needs to be confirmed further.

Usually, neutrophils do not elevate significantly during viral infections. However, viral infection might impair the immune system and facilitate opportunistic infection and neutrophils are crucial in protection against bacterial infections (24). Of interest, the occurrence of sepsis and significantly elevated neutrophils were noticed in patients died with COVID-19, especially in those died within 3 days of admission. This reminded us that secondary infection could be related to the rapid disease progress to death. Frequent detection of bacterial infection indicators like procalcitonin and early implementation of antibiotics might be beneficial for patients suspected of bacterial infections. Compared with patients died after 3 days of admission, prolonged prothrombin time and remarkably increased D-dimer were noticed in patients died within 3 days of admission, which together with depressed platelet count indicated coagulation disturbance and tendency of disseminated intravascular coagulation in these patients.

Up to now, no antiviral treatment has been recommended for coronavirus treatment except supportive care and organ support (6,25). Antiviral medications including remdesivir, lopinavir and traditional Chinese medicine are being used in the clinical practice. Available evidence showed that remdesivir was not associated with statistically signifcant clinical benefts (26). Application of glucocorticoid depended on the severity of inflammatory response. In our study, most nonsurvivors received high flow oxygen inhalation, but quite a low proportion of patients received machine ventilation partly due to limited medical resources against the steeply increasing number of patients in the early outbreak.

Several limitations existed in this study. Firstly, not all laboratory tests were performed in all patients, the missing data might lead to bias and the roles of some indicators might be underestimated. Secondly, in the early outbreak of COVID-19, relatively inadequate medical resources might affect the observation of the natural course of disease. Finally, only 101 patients died with COVID-19 in one hospital were included in this study, the interpretation of our results might be limited, so further studies including multi-centres are still needed.

## CONCLUSIONS

In conclusion, the findings of this study suggested that older patients with comorbidities were at high risk of death with COVID-19. Respiratory failure, acute cardiac and kidney injury played crucial roles in the death of patients. Of importance, the early development of sepsis, increased white blood cell count and neutrophil count indicated poor outcomes and could associated with the rapid disease progress to death. Thus, COVID-19 patients at high risks should attract more attention.

## Data Availability

all original data were saved in Renmin Hospital of Wuhan University, Wuhan, China

## DATAAVAILABILITY STATEMENT

The datasets generated for this study are available on request to these corresponding authors.

## ETHICS STATEMENT

This study was approved by the Ethics Committee of Renmin Hospital of Wuhan University (No. WDRY2020-K059). The ethic committees in this hospital waived informed consent. Oral consent was obtained when we contact family of patients for information of patients’ previous history.

## AUTHOR CONTRIBUTIONS

WW and CC conceptualized the paper. QS, JY and FJ drafted the manuscript. QS, KZ, JY, JF, XZ, XC, PH, YH and ML collected the epidemiological and clinical data. KZ, FJ, KZ contributed to the statistical analyses. JY and FL contributed to radiological analyses. CC and WW were the manuscript’s guarantors and supervised the study. All authors reviewed and approved the final version of the manuscript.

## ACKNOWLEDGMENTS

We thank all the patients and health providers involved in this study.

## References

1. Wang C, Horby PW, Hayden FG, Gao GF. A novel coronavirus outbreak of global health concern. Lancet. (2020) 395: 470–73.

2. World Health Organization. Coronavirus disease 2019 (COVID-19) situation report-105. https://www.who.int/docs/default-source/coronaviruse/situation-reports/20200504-covid-19-sitrep-105.pdf?sfvrsn=4cdda8af2. (accessed May 4 2020).

3. Chang D, Lin M, Wei L, et al. Epidemiologic and clinical characteristics of novel coronavirus infections involving 13 patients outside Wuhan, China. JAMA. (2020) 323: 1092–93.

4. Sanche S, Lin YT, Xu C, Romero-Severson E, Hengartner N, Ke R. High contagiousness and rapid spread of severe acute respiratory syndrome coronavirus 2. Emerg Infect Dis. (2020). https://doi.org/10.3201/eid2607.200282.

5. Chen TM, Rui J, Wang QP, Zhao ZY, Cui JA, Yin L. A mathematical model for simulating the phase-based transmissibility of a novel coronavirus. Infect Dis Poverty. (2020) 9: 24.

6. Wang D, Hu B, Hu C, Zhang F, Liu X, Zhang J, et al. Clinical characteristics of 138 hospitalized patients with 2019 novel coronavirus–infected pneumonia in Wuhan, China. JAMA. (2020) 323: 1061–69.

7. Du Y, Tu L, Zhu P, Mu M, Wang R, Yang P, et al. Clinical features of 85 fatal cases of COVID-19 from Wuhan: A retrospective observational study. Am J Respir Crit Care Med. (2020). https://doi.org/10.1164/rccm.202003-0543OC.

8. Clinical management of severe acute respiratory infection when novel coronavirus (2019-nCoV) infection is suspected: interim guidance, 2020. https://apps.who.int/iris/bitstream/handle/10665/330893/WHO-nCoV-Clinical-2020.3-eng.pdf?sequence=1&isAllowed=y. (accessed January 28 2020).

9. Azoulay E, Mokart D, Kouatchet A, Demoule A, Lemiale V. Acute respiratory failure in immunocompromised adults. The Lancet Respiratory Medicine. (2019) 7: 173–86.

10. ARDS Definition Task Force, Ranieri VM, Rubenfeld GD, Thompson BT, Ferguson ND, Caldwell E, et al. Acute respiratory distress syndrome: The Berlin Definition. JAMA. (2012) 307: 2526–33.

11. Khwaja A. KDIGO clinical practice guidelines for acute kidney injury. Nephron ClinPract. (2012) 120: c179–84.

12. Koch DG, Speiser J, Durkalski V, Fontana RJ, Davern T, McGuire B, et al. The natural history of severe acute liver injury. Am J Gastroenterol. (2017) 112: 1389–96.

13. Singer M, Deutschman CS, Seymour CW, Shankar-Hari M, Annane D, Bauer M, et al. The Third International Consensus Definitions for Sepsis and Septic Shock (Sepsis-3). JAMA. (2016) 315: 801–10.

14. Chung M, Bernheim A, Mei X, Zhang N, Huang M, Zeng X, et al. CT imaging features of 2019 novel coronavirus (2019-nCoV). Radiology. (2020) 295: 202–07.

15. Guan WJ, Ni ZY, Hu Y, Liang WH, Ou CQ, He JX, et al. Clinical Characteristics of Coronavirus Disease 2019 in China. N Engl J Med. (2020) 382: 1708–20.

16. Zhou F, Yu T, Du R, Fan G, Liu Y, Liu Z, et al. Clinical course and risk factors for mortality of adult inpatients with COVID-19 in Wuhan, China: a retrospective cohort study. Lancet. (2020) 395: 1054–62.

17. Jiang F, Deng L, Zhang L, Cai Y, Cheung CW, Xia Z. Review of the Clinical Characteristics of Coronavirus Disease 2019 (COVID-19). J Gen Intern Med. (2020)35:1545–1549.

18. Yang X, Yu Y, Xu J, Shu H, Xia J, Liu H, et al. Clinical course and outcomes of critically ill patients with SARS-CoV-2 pneumonia in Wuhan, China: a single-centered, retrospective, observational study. Lancet Respir Med. (2020) 8: 475–481.

19. Chen T, Wu D, Chen H, Yan W, Yang D, Chen G, et al. Clinical characteristics of 113 deceased patients with coronavirus disease 2019: retrospective study. BMJ. (2020): 368:m1091.

20. Gomersall CD, Joynt GM, Lam P, Li Thomas, Yap F, Lam D, et al. Short-term outcome of critically ill patients with severe acute respiratory syndrome. Intensive Care Med. (2004) 30: 381–87.

21. Santos RAS, Sampaio WO, Alzamora AC, Motta-Santos D, Alenina N, Bader M, et al. The ACE2/angiotensin-(1–7)/ MAS axis of the renin-angiotensin system: focus on angiotensin-(1–7). Physiological reviews. (2018) 98: 505–53.

22. Hashimoto T, Perlot T, Rehman A, Trichereau J, Ishiguro H, Paolino M, et al. ACE2 links amino acid malnutrition to microbial ecology and intestinal inflammation. Nature. (2012) 487: 477–81.

23. Lu R, Zhao X, Li J, Niu P, Yang B, Wu H, et al. Genomic characterisation and epidemiology of 2019 novel coronavirus: implications for virus origins and receptor binding. Lancet. (2020) 395: 565–74.

24. Kolaczkowska E, Kubes P. Neutrophil recruitment and function in health and inflammation. Nat Rev Immunol. (2013) 13: 159–75.

25. de Wit E, van Doremalen N, Falzarano D, Munster VJ. SARS and MERS: recent insights into emerging coronaviruses. Nature Reviews Microbiology. (2016) 14: 523–34.

26. Wang Y, Zhang D, Du G, Du R, Zhao J, Jin Y, et al. Remdesivir in adults with severe COVID-19: a randomised, double-blind, placebo-controlled, multicentre trial. Lancet. (2020). https://doi.org/10.1016/S0140-6736(20)31022-9.

